# SARS-CoV-2 antibody prevalence among homeless people, sex workers and shelter workers in Denmark: a nationwide cross-sectional study

**DOI:** 10.1101/2021.05.07.21256388

**Authors:** Alexandra R Röthlin Eriksen, Kamille Fogh, Rasmus B Hasselbalch, Henning Bundgaard, Susanne D Nielsen, Charlotte S Jørgensen, Bibi FSS Scharff, Christian Erikstrup, Susanne G. Sækmose, Dorte K Holm, Bitten Aagaard, Jonas H Kristensen, Cecilie A Bødker, Jakob Norsk, Pernille Brok Nielsen, Lars Østergaard, Svend Ellermann-Eriksen, Berit Andersen, Henrik Nielsen, Isik S. Johansen, Lothar Wiese, Lone Simonsen, Thea K.Fischer, Fredrik Folke, Freddy Lippert, Sisse R Ostrowski, Steen Ethelberg, Anders Koch, Anne-Marie Vangsted, Tyra Krause, Anders Fomsgaard, Claus Nielsen, Henrik Ullum, Robert Skov, Kasper Iversen

## Abstract

**Background:** People experiencing homelessness (PEH) and associated shelter workers may be at higher risk of infection with “Severe acute respiratory syndrome coronavirus 2” (SARS-CoV-2). The aim of this study was to determine the prevalence of SARS-CoV-2 among PEH and shelter workers in Denmark.

**Design and methods:** In November 2020, we conducted a nationwide cross-sectional seroprevalence study among PEH and shelter workers at 21 recruitment sites in Denmark. The assessment included a point-of-care test for antibodies against SARS-CoV-2, followed by a questionnaire. The seroprevalence was compared to that of geographically matched blood donors considered as a proxy for the background population, tested using a total Ig ELISA assay.

**Results:** We included 827 participants in the study, of whom 819 provided their SARS-CoV-2 antibody results. Of those, 628 were PEH (median age 50.8 (IQR 40.9-59.1) years, 35.5% female) and 191 were shelter workers (median age 46.6 (IQR 36.1-55.0) years and 74.5% female). The overall seroprevalence was 6.7% and was similar among PEH and shelter workers (6.8% vs 6.3%, p=0.87); and 12.2% among all participants who engaged in sex work. The overall participant seroprevalence was significantly higher than that of the background population (2.9%, p <0.001). When combining all participants who reported sex work or were recruited at designated safe havens, we found a significantly increased risk of seropositivity compared to other participants (RR 2.1, 95% CI 1.16-3.75, p=0.02). Seropositive and seronegative participants reported a similar presence of at least one SARS-CoV-2 associated symptom (49% and 54%, respectively).

**Interpretations:** The prevalence of SARS-CoV-2 antibodies was more than twice as high among PEH and associated shelter workers, compared to the background population. The subset of the study participants who were also sex workers were at particularly high risk of COVID-19 infection.

**Funding:** TrygFonden and HelseFonden.

## Introduction

Severe acute respiratory syndrome coronavirus 2 (SARS-CoV-2), has since its emergence in China in 2019 caused a global pandemic. As of April 26^th^ 2021 an estimated 146 million people worldwide was infected and more than 3 million people has died from SARS-CoV-2 (1). The first confirmed case of SARS-CoV-2 in Denmark was detected on February 27^th^ 2020 and since then there has been more than 246 460 confirmed cases of SARS-CoV-2 in Denmark (2). Vulnerable groups including people experiencing homelessness (PEH) have challenges in accessing health care systems and public health information (3). Limited knowledge of protection against SARS-CoV-2 among vulnerable individuals such as PEH is likely to increase the risk of infection for both PEH and people in their proximity, such as shelter workers. Additionally, the recommended guidelines to prevent the spread of SARS-CoV-2 might not be feasible due to inadequate access to handwash, protective equipment and difficulties in practicing social distancing (4,5). An estimated 6,431 (0.1%) Danes are categorized as homeless, of whom a total of 2.666 (41.5%) are registered in the Capital Region of Denmark and the rest are distributed throughout the largest cities in Denmark (6,7). Crowded living conditions in shelters and public spaces where PEH reside, constitute a potential risk of becoming epicenters, as congregate settings have proven to be associated to high exposure to SARS-CoV-2 (8). Furthermore, PEH have more physical and mental health issues than the background population (9,10) and often engage in substance abuse (6), which could further increase their risk of infection and of a serious course of disease by SARS-CoV-2 (11,12). A fear of experiencing serious withdrawal symptoms may prevent this group from both testing and subsequent self-isolation. Some PEH engage in sex work which may further increase the risk of infection with SARS-CoV-2, due to direct physical contact with clients (13). Sex workers are known to have a high prevalence of HIV (14) and other underlying health conditions (15,16), which may add to their risk of SARS-CoV-2 progressing to severe illness (12). Systematic screening for SARS-CoV-2 antibodies is an important tool in the surveillance of the current pandemic. A French study found the overall seroprevalence among PEH to be 52.1% which was 4.3 times higher than the modelled estimate for the general population in Ile de France (12%) (8). Information on the prevalence of SARS-CoV-2 infection among vulnerable groups such as PEH is important to assess the need for preventive measures in such groups, to provide information about support estimations of the overall prevalence of SARS-CoV-2 infection and to help guide the public health response in the future. This study is part of the national surveillance study “Testing Denmark”, aimed at assessing the extent and impact of SARS-CoV-2 infection in Denmark (17). The aim of the present study was to determine SARS-CoV-2 seropositivity among PEH and shelter workers in Denmark, and to study risk factors for infection and clinical presentation in PEH.

## Design and methods

### Study design and sampling strategy

We conducted a nationwide, cross-sectional seroprevalence study between November 2^nd^ and 20^th^ 2020, to determine the prevalence of SARS-CoV-2 antibodies among PEH using a rapid point-of care SARS-CoV-2 antibody test (POCT). In addition, we also included shelter workers at the recruitment sites. Participants were invited to fill in a questionnaire in collaboration with a trained project employee, at the same time as the test was performed. We recruited participants from 21 sites in the four biggest cities in Denmark; Copenhagen, Aarhus, Aalborg and Odense. The recruitment sites were shelters, supervised sites for intravenous drug abusers, food distribution sites, meeting places and day/night cafés. In the week prior to our visit, written information was distributed by shelter workers at the recruitment sites notifying the participants of our project. To ensure a high attendance and inclusion, recruitment sites were visited several times, on different days and at different time of the day, including weekends and evenings. Participants were encouraged to wait around for15 minutes for their test results, but in case they did not want to, they were contacted if the test result was positive. Most came back throughout the day to receive their test result.

### SARS-CoV-2 in Denmark

The first confirmed case of SARS-CoV-2 in Denmark was detected on February 27^th^, 2020. Since then, there have been more than 246 460 confirmed cases of SARS-CoV-2 in Denmark (2). Overall, more than 12 880 of the Danish SARS-CoV-2 patients were hospitalized between January 2020 and March 2021.

### Background population

All Danish blood donations are routinely screened for the presence of SARS-CoV-2 antibodies since October 2020. Blood donations take place in all five Danish administrative regions (17) and donors are 17-69 years old. The seroprevalence estimates from the period between the 1^st^ and 22^nd^ of November 2020, are used in this study as proxies for the SARS-CoV-2 infection in the background population.

### Point of care test

SARS-CoV-2 antibodies in PEH and shelter workers were detected in whole blood, by use of the OnSite COVID-19 IgG/IgM Rapid Test (CTK Biotech inc., Poway, California, United States of America) according to the manufacturer’s recommendations. This POCT is a single use lateral flow chromatographic immunoassay rapid test, intended for qualitative detection and differentiation of SARS-CoV-2 immunoglobulin M (IgM) and immunoglobulin G (IgG) antibodies. Blood was extracted from the fingertip. Test results were read after 15 minutes by a trained project employee. Participants were categorized as seropositive if they were either IgG and/or IgM positive. The test sensitivity and specificity is 96.86% (95% confidence interval (95% CI:93.66%-98.47%) and 99.39% (95% CI 97.80%-99.83%) respectively(18), as reported from the manufacturer. In house validation of CTK’s POCT showed a slightly lower sensitivity of 90% and a specificity of 100% (19). In contrast, blood donors were screened for seropositivity using an Enzyme-Linked Immunosorbent Assay (ELISA), with a sensitivity of 96.7% (95% CI 92.4-98.6) and specificity of 99.5% (95% CI 98.7-99.8) (Beijing Wantai Biological Pharmacy Enterprise, China).

### Questionnaire

Participants were invited to fill in a questionnaire provided at the recruitment site, comprised of questions covering sociodemographic characteristics, physical health, use of drugs and alcohol, co-morbidities, symptom manifestations and the use of personal protective equipment against SARS-CoV-2. Alcohol abuse was defined as drinking more than the national recommendations of <1 or <2 beverages per day for women and men, respectively (22). The questionnaire was filled out with the assistance of a trained project employee. Personal data was collected using a web-based electronic data capture tool (Research Electronic Data Capture, REDCap) (20,21).

### Study group

Homelessness was defined as people living rough, in emergency accommodation, in accommodation for the homeless and living in non-conventional dwellings due to lack of housing according to the ETHOS (European Typology on Homelessness and Housing Exclusion) classification (23) established by the FEANTSA (European Federation of National Organizations Working with the Homeless) (24). Sex workers were defined as individuals either having reported to be engaged in sex work in the questionnaire and/or being included at one of the designated safe havens for sex workers; Reden and Reden International. Shelter workers are defined as people who either work or volunteer at the recruitment sites.

### Primary outcome

The primary outcome was the proportion of the study population with a positive antibody test (IgG and/or IgM) for SARS-CoV-2.

### Ethics

This study was performed as a national surveillance study under the authority task of Statens Serum Institut (SSI; Copenhagen, Denmark, the Danish National Institute for Infectious Disease Control and Prevention which performs the epidemiological surveillance of infectious diseases for the Danish government), hence does not require any formal approval from an ethics committee according to Danish law. This decision was made by the regional Ethics Committee of the Capital Region in Denmark (20057075). This study was carried out in agreement with the Helsinki II declaration. All participation in this study was voluntary. This study was registered with the Danish Data Protection Authorities (P-2020-901). All personal data, obtained in REDCap, was kept in accordance with the general data protection regulation and data protection law stated by the Danish Data Protection Agency.

### Statistical analysis

Seropositivity is presented as numbers (n) and percentage (%) with 95% confidence intervals (95% CI). Baseline characteristics and exposures are presented as n (%) for factors and median and interquartile range (IQR)) for numeric variables as appropriate. Answers with “do not know” were classified as missing and answers marked “not relevant” were classified as “no”. We tested for significance using students T-test, Wilcoxon rank sum test or Fischer’s exact test. Significant risk factors of seropositivity were combined in a multivariate logistic regression model including region. P-values <0.05 was considered significant. Analyses were performed using RStudio version 1.2.5001 (25).

## Results

### Characteristics

We recruited a total of 827 participants between the 2^nd^ and 20^th^ of November 2020 of which 819 provided their SARS-CoV-2 antibody result as illustrated in figure 1. Participants were recruited from 21 recruitment sites placed in Copenhagen (n = 351), Odense (n = 128), Aarhus (n = 144) and Aalborg (n = 144). Further, 52 participants did not register their test location. The recruited participants included 628 PEH (median age 50.8 (IQR 40.9-59.1) years, 35.5% female) and 191 shelter workers (median age 46.6 (IQR 36.1-55.0) years, 74.5% female). Baseline characteristics of the cohort is shown in table 1. The PEH were older (p<0.001), had a lower BMI (p=0.02) and were more likely to be male (p<0.001) and smoke (p<0.001). Supplementary table 1 shows baseline characteristics of all participants stratified according to seropositivity. The only significant difference was for BMI which was significantly higher among the seropositive group (<0.001).

**Figure 1:**
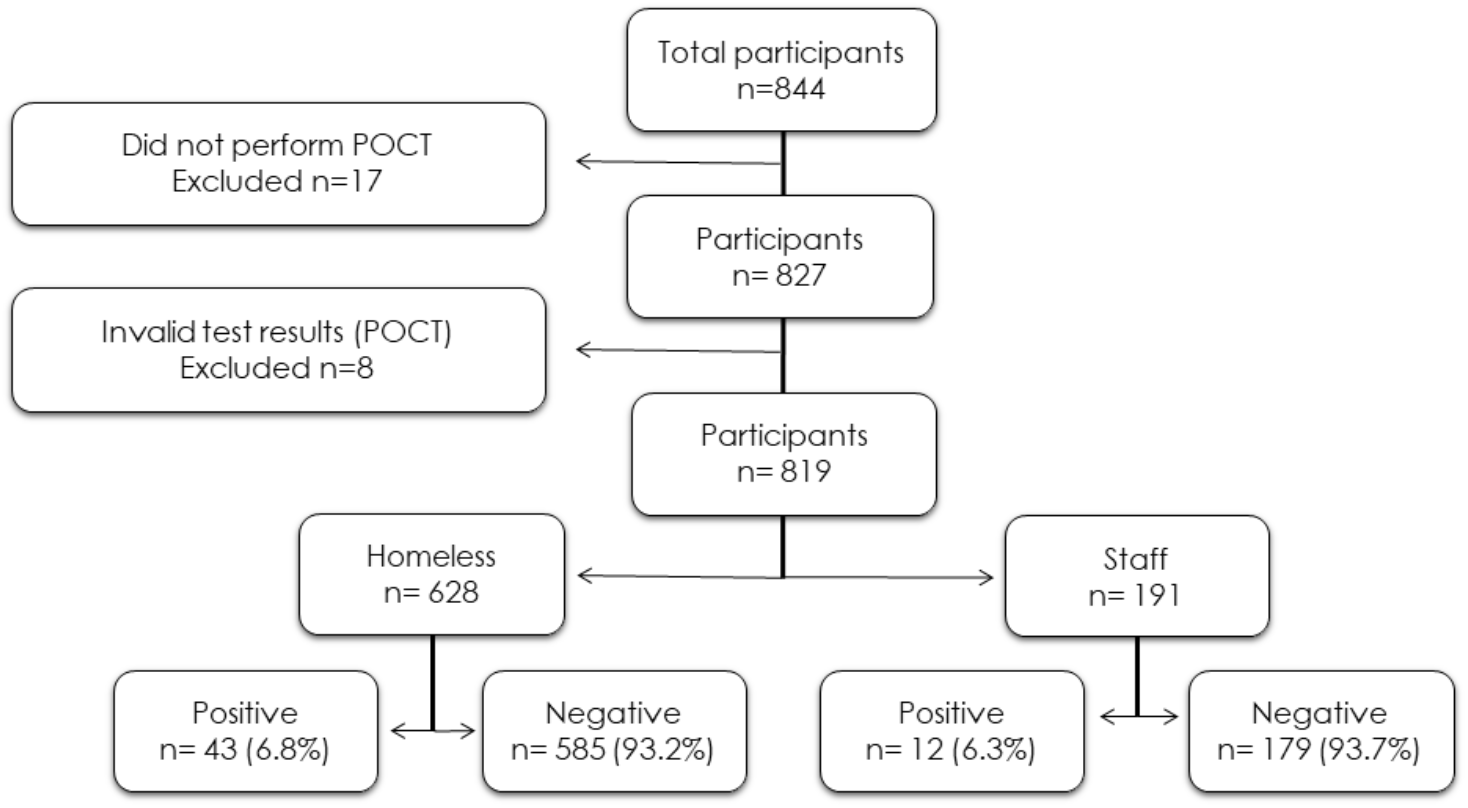
Flow chart reporting the flow of participants through each stage.

**Table 1:** Baseline Characteristics of the study cohort of shelter workers and people experiencing homelessness (PEH).

|  | PEH | Shelter Workers | p |
| --- | --- | --- | --- |
| **n** | 628 | 191 |  |
| **Seropositive (%)** | 43 (6.8) | 12 (6.3) | 0.914 |
| **Age (median [IQR])** | 50.83 [40.86, 59.14] | 46.62 [36.09, 54.99] | <0.001 |
| **Gender (%)** |  |  | <0.001 |
| Female | 219 (34.9) | 140 (73.3) |  |
| Male | 396 (63.1) | 48 (25.1) |  |
| Other | 2 (0.3) | 0 (0.0) |  |
| NA | 11 (1.8) | 3 (1.6) |  |
| **Body mass index (mean (SD))** | 24.75 (5.02) | 25.92 (5.42) | 0.024 |
| **Smoker (%)** | 498 (79.3) | 89 (46.6) | <0.001 |
| **Alcohol use (%)** |  |  | <0.001 |
| Yes | 307 (48.9) | 166 (86.9) |  |
| NA | 32 (5.1) | 5 (2.6) |  |
| **Previously tested (%)** |  |  | <0.001 |
| Yes | 371 (59.1) | 154 (80.6) |  |
| NA | 22 (3.5) | 3 (1.6) |  |

PEH: people experiencing homelessness. Seropositive: SARS-CoV-2 IgM and/or IgG antibodies detected in POCT. Smoker: either previously or currently smoking. Alcohol use: intake of alcohol within the past 12 months. Previous SARS-CoV-2 Tested: have previously been tested for SARS-CoV-2

### Seroprevalence

Of the 819 participants, 55 (6.7%) were seropositive. We found that 43 of 628 (6.8%) PEH and 12 of 191 (6.3%) shelter workers were seropositive, the prevalence in the two groups was not significantly different (p=0.87).

### Seroprevalence compared to the background population

In the period between the 1^st^ and 22^nd^ of November 2020, the background population (n= 18505) had a 2.9% seroprevalence. This group was characterized by a median age of 43 (IQR 29-54) years and a higher proportion (47.9%) were women. Taken together, the participants in our study (PEH and shelter workers combined) were at a significantly higher risk of seropositivity than the background population (RR 2.2, 95% CI 1.75-2.99, p<0.001). Figure 2 illustrates the regional SARS-CoV-2 seroprevalence in PEH, shelter workers and the background population.

**Figure 2:**
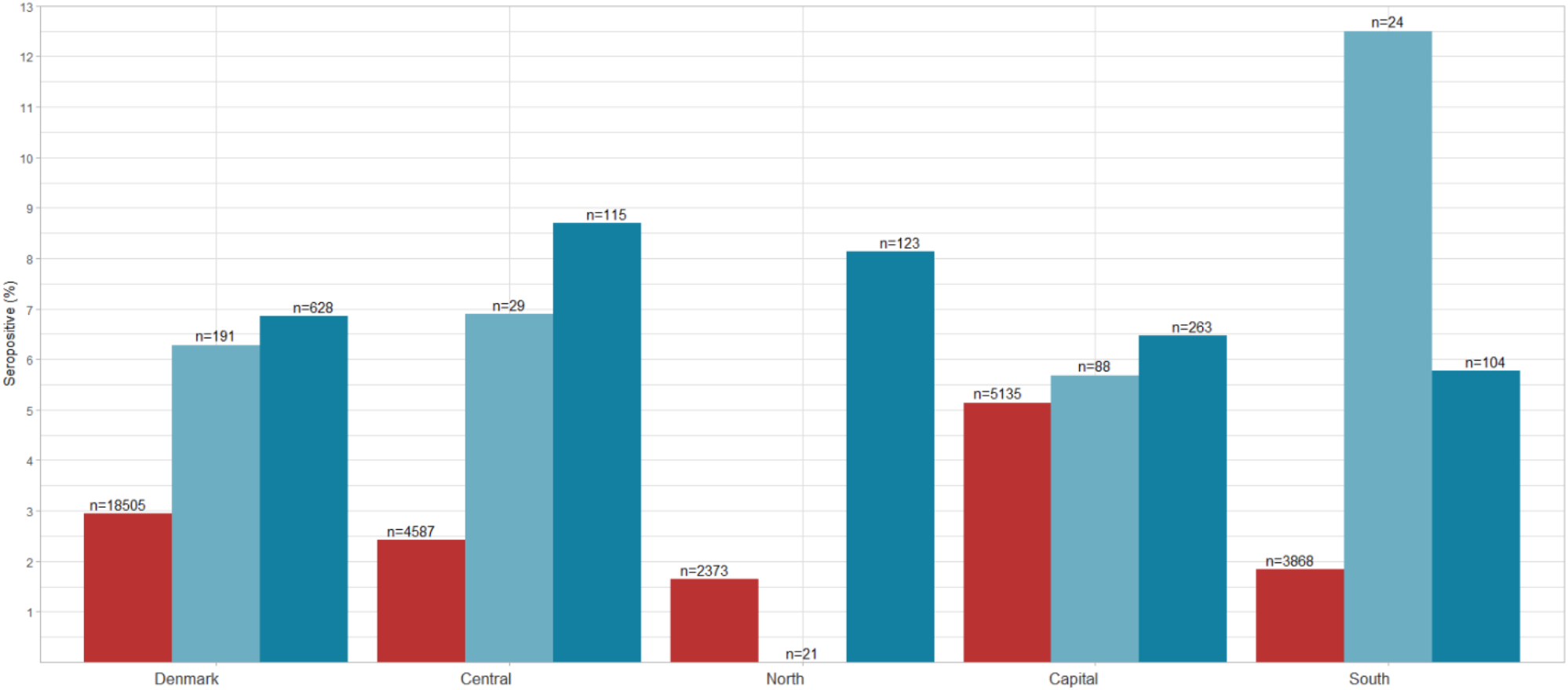
Seroprevalence among PEH and shelter workers, compared to the background population. Red: blood donors serving as proxy for the general population; Light blue: shelter workers; Dark blue: people experiencing homelessness.

The seroprevalence among PEH alone was also significantly higher than that in the background population (RR 2.3, 95% CI 1.73-3.14, p <0.001).

The subset of shelter workers (n=191) in general was significantly associated with seropositivity compared to the background population (RR 2.1, 95% CI 1.23-3.72, p=0.02).

### Risk factors for attracting infection

Table 2 shows the prevalence of risk factors (drugs, alcohol, sex work) in the study population, stratified by seropositivity. Although sex workers were 1.9 times more likely to be seropositive compared to non sex workers (OR 1.9), none of the lifestyle risk factors were significantly associated to seropositivity.

**Table 2:** Characteristics and risk factors stratified according to seropositivity stratified by PEH and shelter worker (IgM and/or IgG).

|  | Seropositive PEH | Seronegative PEH | p |
| --- | --- | --- | --- |
| **n** | 43 | 585 |  |
| **Drug abuse (%)** | 9 (20.9) | 221 (37.8) | 0.040 |
| IV drugs (%) | 8 (18.6) | 154 (26.3) | 0.349 |
| Smoked drugs (%) | 11 (25.6) | 192 (32.8) | 0.418 |
| **Alcohol use (%)** |  |  | 0.019 |
| Yes | 21 (48.8) | 286 (48.9) |  |
| NA | 6 (14.0) | 26 (4.4) |  |
| **Alcohol abuse (%)** |  |  | 0.549 |
| Yes | 8 (18.6) | 132 (22.6) |  |
| NA | 28 (65.1) | 331 (56.6) |  |
| **Sex work (%)** | 8 (18.6) | 64 (10.9) | 0.202 |
| **Contact with known SARS-CoV-2 infected person:** |  |  |  |
| **15min Contact (%)** |  |  | 0.550 |
| Yes | 4 (9.3) | 45 (7.7) |  |
| NA | 37 (86.0) | 527 (90.1) |  |
| **Physical Contact (%)** |  |  | 0.377 |
| yes | 2 (4.7) | 14 (2.4) |  |
| NA | 36 (83.7) | 528 (90.3) |  |
| **Social Circle (%)** |  |  | 0.443 |
| Yes | 8 (18.6) | 70 (12.0) |  |
| NA | 7 (16.3) | 100 (17.1) |  |
|  | Seropositive Shelter Workers | Seronegative Shelter Workers | p |
| **n** | 12 | 179 |  |
| **Drug abuse (%)** | 0 (0.0) | 3 (1.7) | 1.000 |
| IV drugs (%) | 0 (0.0) | 0 (0.0) |  |
| Smoked drugs (%) | 0 (0.0) | 3 (1.7) | 1.000 |
| **Alcohol use (%)** |  |  | 0.809 |
| Yes | 11 (91.7) | 155 (86.6) |  |
| NA | 0 (0.0) | 5 (2.8) |  |
| **Alcohol abuse (%)** |  |  | 0.381 |
| Yes | 2 (16.7) | 14 (7.8) |  |
| NA | 1 (8.3) | 37 (20.7) |  |
| **Sex work (%)** | 1 (8.3) | 1 (0.6) | 0.273 |
| **Contact with known SARS-CoV-2 infected person:** |  |  |  |
| **15min Contact (%)** |  |  | 0.640 |
| Yes | 1 (8.3) | 27 (15.1) |  |
| NA | 11 (91.7) | 146 (81.6) |  |
| **Physical Contact (%)** |  |  | 0.451 |
| Yes | 1 (8.3) | 12 (6.7) |  |
| NA | 11 (91.7) | 146 (81.6) |  |
| **Social Circle (%)** |  |  | 0.423 |
| Yes | 2 (16.7) | 60 (33.5) |  |
| NA | 4 (33.3) | 39 (21.8) |  |

PEH: people experiencing homelessness. IV drugs: use of intravenous drugs. Smoked drugs: drugs that were reported smoked. Alcohol use: intake of alcohol within the past 12 months. Alcohol Abuse: drinking more than the national recommendations of 2 drinks or less in a day for men or 1 drink or less in a day for women. 15 min. contact: having been in the same room as someone with an ongoing SARS-CoV-2 infection for 15 minutes or longer. Physical contact: having had close bodily contact with a SARS-CoV-2 positive individual. Social circle: aware of anyone in close contacts who had SARS-CoV-2.

A total of 285 (45.4%) PEH reported use of drugs, the most commonly used were cocaine (18.6%) and heroin (15.6%). Among the seropositive PEH, heroin (14.5%) was the most common drug while the seronegative PEH were more likely to use cocaine (16.9%). Further, we found that 326 (51.9%) PEH were smokers; of those 326 (51.9%) smoked tobacco and 180 (28.7%) smoked cannabinoids. Other reported smoked substances were cocaine (3.5%) and heroin (2.5%). A total of 307 (51.5%) PEH reported some use of alcohol within the past year.

Of the 191 shelter workers only 3 (1.6%) reported using drugs, all of which were of cannabinoids. Further, 166 (89.2%) reported any use of alcohol within the past year. PEH were significantly more likely to abuse alcohol (p<0.001) and use drugs (p<0.001), compared to the shelter staff.

### Sex work

Of the 72 (11.5%) PEH who engaged in sex work, 8 (11.1%) were seropositive. Table 3 illustrates characteristics and risk factors of PEH stratified by sex work. Sex workers were younger, more likely to be female and less likely to report smoking and/or IV drug use than other PEH (all p<0.001). Further, two (1.0%) shelter workers reported having engaged in sex work, of whom one was found to be SARS-CoV-2 seropositive. The seroprevalence among all participants, both PEH and shelter workers, engaging in sex work (n =74) was 12.2%.

**Table 3:** Characteristics and risk factors for people experiencing homelessness (PEH), stratified by engagement in sex work.

|  | Sex Workers | PEH not engaging in sex work | p |
| --- | --- | --- | --- |
| **n** | 72 | 556 |  |
| **Seropositive (%)** | 8 (11.1) | 35 (6.3) | 0.202 |
| **Age (median [IQR])** | 41.43 [32.51, 49.96] | 51.57 [41.64, 59.52] | <0.001 |
| **Gender (%)** |  |  | <0.001 |
| Female | 68 (94.4) | 151 (27.2) |  |
| Male | 3 (4.2) | 393 (70.7) |  |
| Other | 1 (1.4) | 1 (0.2) |  |
| NA | 0 (0.0) | 11 (2.0) |  |
| **Smoker (%)** | 44 (61.1) | 454 (81.7) | <0.001 |
| **Drug abuse (%)** | 17 (23.6) | 213 (38.3) | 0.021 |
| IV Drugs (%) | 6 (8.3) | 156 (28.1) | 0.001 |
| Smoked drugs (%) | 19 (26.4) | 184 (33.1) | 0.312 |
| **Alcohol use (%)** |  |  |  |
| Yes | 27 (37.5) | 280 (50.4) | <0.001 |
| NA | 16 (22.2) | 16 (2.9) |  |
| **Alcohol abuse (%)** |  |  |  |
| Yes | 10 (13.9) | 130 (23.4) | 0.172 |
| NA | 47 (65.3) | 312 (56.1) |  |
| **Contact with known SARS-CoV-2 infected person:** |  |  |  |
| **15 Min Contact (%)** |  |  |  |
| Yes | 3 (4.2) | 46 (8.3) | 0.467 |
| NA | 67 (93.1) | 497 (89.4) |  |
| **Body Contact (%)** |  |  |  |
| Yes | 1 (1.4) | 15 (2.7) | 0.610 |
| NA | 67 (93.1) | 497 (89.4) |  |
| **Social Circle (%)** |  |  |  |
| Yes | 8 (11.1) | 70 (12.6) | 0.036 |
| NA | 20 (27.8) | 87 (15.6) |  |
| **Previously tested (%)** |  |  |  |
| Yes | 39 (54.2) | 332 (59.7) | <0.001 |
| NA | 15 (20.8) | 7 (1.3) |  |

Sex workers: PEH engaging in sex work. PEH: People experiencing homelessness. Seropositive: SARS-CoV-2 IgM and/or IgG antibodies detected in POCT. Smoker: either previously or currently smoking. IV drugs: use of intravenous drugs. Smoked drugs: drugs that were reported smoked. Alcohol use: intake of alcohol within the past 12 months. Alcohol Abuse: drinking more than the national recommendations of 2 drinks or less in a day for men or 1 drink or less in a day for women. Never COVID-19 Tested: never previously been tested for COVID-19.

For all participants engaging in sex work (n=74), there was a significantly increased risk of seropositivity for IgG antibodies compared to the rest of the study group (RR 2.8, 95% CI 1.26-6.29, p=0.02). However, for the combined seropositivity (IgG and/or IgM antibodies) the difference did not reach significance (RR 2.0, 95% CI 1.00-3.86, p=0.08).

We included a total of 33 shelter workers working at designated safe havens for sex workers, of whom 4 (12.1%) were found to be seropositive. Shelter workers at designated safe havens for sex workers had a far greater infection rate than other shelter workers, however, the difference was not significant (RR 2.4, 95% CI 0.53-10.38, p=0.13). Further, there was no difference in seropositivity between shelter workers at designated safe havens (12.1%) and all sex workers (12.2%) (RR 1.3, 95% CI 0.47-3.75, p=0.75). When combining all participants who reported sex work or were recruited at designated safe havens, shelter workers and sex workers (n=107), we found a significantly increased risk of seropositivity (IgG and/or IgM antibodies) compared to other participants (RR 2.1, 95% CI 1.16-3.75, p=0.02). In a multivariate logistic regression model of region of stay, being a sex worker or working at a designated safe haven remained a significant risk factor of seropositivity compared to PEH who does not engage in sex work (OR 2.4, 95% CI 1.12-4.70, p=0.02).

### Symptoms and self-reported illness

Of the 628 PEH, 57 (9.1%) suspected previous infection with SARS-CoV-2 of whom 8 (14%) were seropositive. A total of 371 (59.1%) PEH reported being previous tested for SARS-CoV-2, of whom 11 (3.0%) were reportedly positive at the time. However, of those only 3 (27.3%) tested positive for SARS-CoV-2 antibodies in our study. Of the shelter workers, 154 (80.6 %) reported previous testing for SARS-CoV-2. No shelter workers reported having previously tested positive for SARS-CoV-2, but 21 (11%) suspected previous infection with SARS-CoV-2 of whom 5 (23.8%) were seropositive.

Seventeen of 43 (39.5%) seropositive PEH and 10 of the 12 (83.3%) seropositive shelter workers reported having had symptoms. Overall, a total of 441 (53.8%) participants in our study group reported having had one or more symptoms. Of those, 15 (3.3%) reported being hospitalized at the time of their symptoms. Among the PEH 303 (48.2%) reported experiencing one or more symptom, of whom 26 (8.6%) though it was attributable to SARS-CoV-2. Of the shelter workers 138 (72.3%) reported one or more symptom, and of those 20 (14.5%) thought the symptoms were attributable to SARS-CoV-2; significantly more than among the PEH (RR 1.7, 95%CI: 1.36-2.22, p<0.001). Similarly, shelter workers were more likely than PEH to have more than three symptoms (RR 1.7, 95%CI: 1.36-2.22, p<0.001). The most common symptoms reported in the combined cohort were fever (23.2%) and shivers (18.5%). However, there was no observed significant association between experiencing symptoms and seropositivity (RR 0.8, 95%CI: 0.50-1.38, p=0.49). Table 4 shows symptoms stratified by seropositivity. Symptoms in PEH and shelter workers, is illustrated in supplementary table 2.

**Table 4:** Symptoms reported by the participants, stratified by serology findings.

|  | Seropositive | Seronegative | p |
| --- | --- | --- | --- |
| **n** | **55** | **764** |  |
| **Any symptom (%)** | 27 (49.1) | 414 (54.2) | 0.554 |
| **Fever $\geq 38^{\circ}\text{C}$ (%)** | 8 (14.5) | 182 (23.8) | 0.159 |
| **Chills (%)** | 10 (18.2) | 142 (18.6) | 1.000 |
| **Loss of Smell (%)** | 8 (14.5) | 63 (8.2) | 0.175 |
| **Loss of Taste (%)** | 7 (12.7) | 54 (7.1) | 0.201 |
| **Sore Throat (%)** | 12 (21.8) | 238 (31.2) | 0.194 |
| **Cough (%)** | 16 (29.1) | 315 (41.2) | 0.103 |
| **Shortness of breath (%)** | 8 (14.5) | 154 (20.2) | 0.404 |
| **$\geq 3$ Symptoms (%)** | 11 (20.0) | 204 (26.7) | 0.351 |

Symptoms experienced since March 1<sup>st</sup>, 2020. $\geq 3$ Symptoms; participants who registered three or more symptoms.

### Use of protective means against SARS-CoV-2 infection

Figure 3 shows use of protective means against SARS-CoV-2 in PEH compared to shelter workers. Only 25 (4%) of the PEH reported that they did not follow any of the recommended guidelines. Among the shelter workers only 3 (1.6%) reported not following the guidelines.

**Figure 3:**
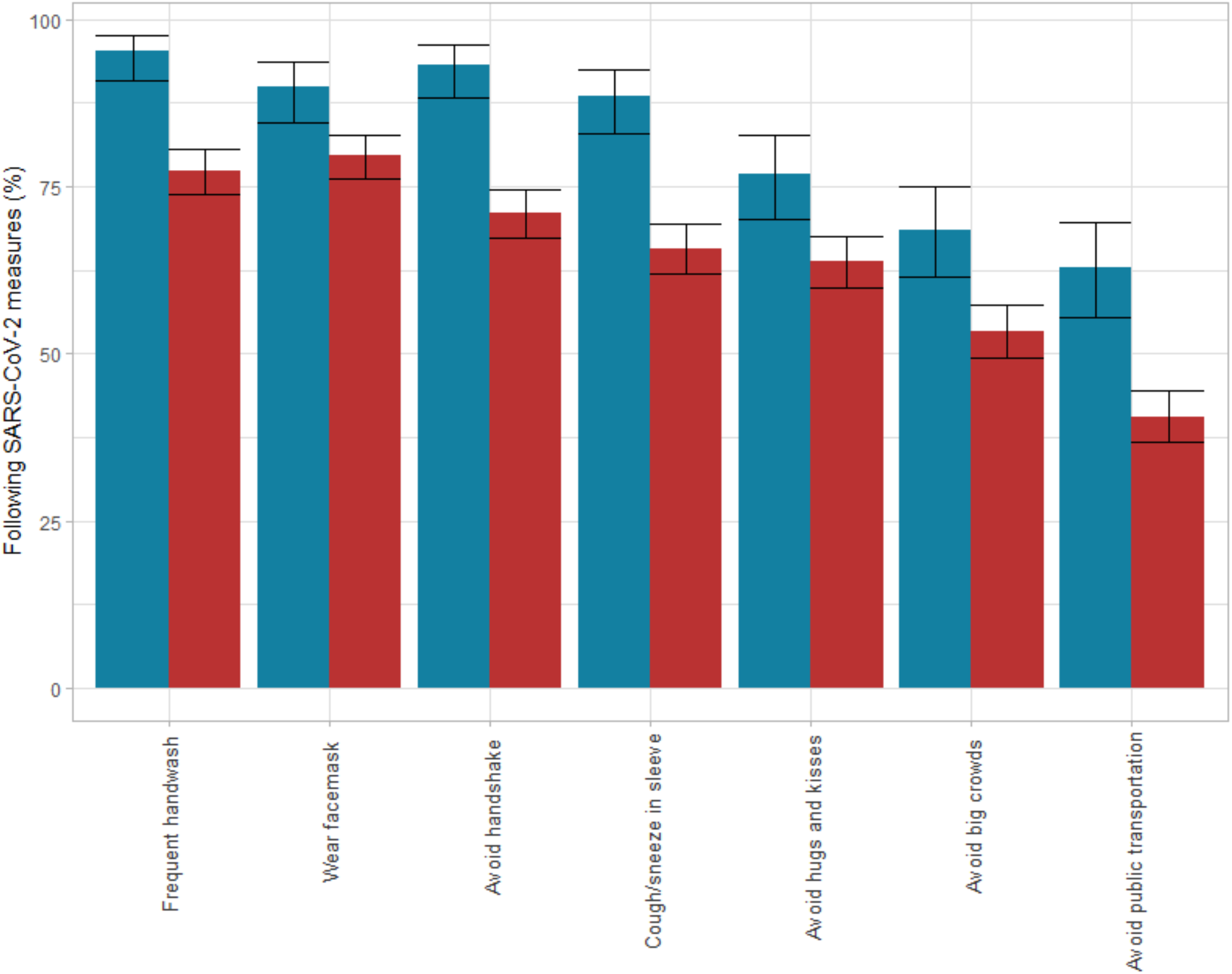
Percentage of PEH and shelter workers who follow the national SARS-CoV-2 measures and guidelines. Blue: Shelter workers; Red: people experiencing homelessness (PEH). 95% CI is illustrated.

## Discussion

To our knowledge, our study is the first to investigate and evaluate the nationwide prevalence of SARS-CoV-2 infection among PEH. We found that the seroprevalence among PEH was twice that of the background population. Furthermore, sex workers and shelter workers at designated safe havens were at increased risk of infection with SARS-CoV-2, independent of region. We found that seropositive PEH were less likely to report symptoms, compared to seropositive shelter workers. The results further suggest that almost all PEH follow one or more national SARS-CoV-2 prevention measure, as illustrated in figure 3. The high seroprevalence among PEH found in our study could be taken into consideration when deciding in which phase they are eligible for a vaccine, as part of the national SARS-CoV-2 vaccination program rollout.

Our findings are consistent with findings in previous studies(8,26–28), with elevated prevalence of SARS-CoV-2 for people living in precarious conditions, relative to the background population. This is consistent with living in overcrowded conditions, a main risk factor associated with infection of SARS-CoV-2.

The observed increased seroprevalence in sex workers compared to those who did not engage in sex work, is in accordance with the recent statement from UNAIDS (the joint United Nations Programme on HIV/AIDS) who emphasized how sex workers are risking their health by working during the current SARS-CoV-2 pandemic in order to provide for themselves (29). Further, we believe that national SARS-CoV-2 measures such as social distancing are simply not feasible for sex workers as their work requires some level of close physical contact with clients and self-isolation could result in a total loss of income. These results show that the risk of contracting SARS-CoV-2 should be added to the risks experienced by sex workers.

In previous studies around half the seropositive participants are reporting symptoms attributable to SARS-CoV-2 (30–32). Symptom prevalence in our study is consistent with previous findings on PEH and implies a high proportion of asymptomatic infections (8,26,33,34). Additional explanations might include difficulties in recalling previous symptoms and differentiating symptoms attributable to substance abuse and SARS-CoV-2. Thus, symptom assessment in PEH might not be predictive for SARS-CoV-2. The high prevalence of substance abuse among PEH is consistent with previous national findings on PEH (6). Recent studies suggest that suffering from a substance use disorder increases the risk of contracting SARS-CoV-2, while also facing a worse outcome than the background population (3,35).

### Strengths and Limitations

Our study has several limitations. First, the cross-sectional study designs do not allow determination of time of infection nor provide information on when participants became seropositive. Individuals who might already have tested positive by PCR at an earlier point in time, might have chosen not to participate in this study. Individuals anticipating a positive result might have chosen not to participate, fearful of the consequences such as isolation. If so, our results could be biased and the seroprevalence be underestimated. However, our apprehension is that the desire to participate and get tested was high (10.2% of an estimated 6 431 homeless people in Denmark). Of the 827 participants in our study group, 544 (67.0%) reported having previously been tested (nasopharyngeal swab and/or antibody test) and still participated. Further, questions on sociodemographic characteristics, physical health, use of drugs and alcohol, co-morbidities, symptom manifestations and the use of personal protective equipment against SARS-CoV-2 were self-reported, hence an information bias could have affected our results We compared our seropositivity findings to that of blood donors serving as a proxy to the background population, with some limitations to consider. First, blood donors are required to have a good general health and are ineligible to donate blood if they have ever engaged in sex work, have certain medical conditions, travel to certain international destinations and/or receive certain immunizations. Seropositivity could as a result potentially be lower in this group of the population compared to the background population. Although, blood donors due to their good health, are more represented in the labor market and so more at risk of SARS-CoV-2 infection. Another limitation is that blood donors and our study group were tested using different methods, POCT vs ELISA however both with high and similar sensitivity and specificity. Antibodies generated in response to SARS-CoV-2 exposure are generated in the weeks after the acute phase of the infection, thus it may not register in recently infected participants. However, recent studies have shown that SARS-CoV-2 IgM antibodies reach threshold to be detected 5-7 days after symptom onset (25). Thus, participants currently or recently infected with SARS-CoV-2 might not have been identified in this study. A strength of serology approach versus test by PCR is that it allows us to detect SARS-CoV-2 antibodies in those categorized as asymptomatic carriers and those with suspected SARS-CoV-2 despite negative PCR results. Furthermore, serology testing with a POCT is easy and can be performed as a self-test. It does not require a blood sample nor laboratory equipment, hence is less costly than a laboratory serology test and provide more rapid results.

## Conclusions

In this study we found a high SARS-CoV-2 seroprevalence among PEH and shelter workers, compared to the background population. Study participants who reported sex work were at a fourfold elevated risk of being SARS-CoV-2 seropositive. There was no significant association between reported symptoms and seropositivity, nor between substance abuse and seropositivity.

## Data Availability

No data are available

## Author contributions

The study was designed and initiated by Kasper Iversen, Robert Skov and Kamille Fogh. Data was collected by: Alexandra R Röthlin Eriksen and Kamille Fogh. Data analysis was done by Alexandra R Röthlin Eriksen and Rasmus Hasselbalch. The first draft was written by Alexandra R Röthlin Eriksen, Kamille Fogh, Rasmus Hasselbalch, Henning Bundgaard and Kasper Iversen

All authors have critically revised the manuscript and agree to be accountable for all aspects of the work. All authors approved the final version of the manuscript.

## Acknowledgements

The authors would like to thank Kåre Mølbak, Thomas Benfield and Cyril Martel for their contributions to this study. The authors would also like to thank Mændenes Hjem, Café Klare, Herbergscentret Sundholm, The nest international, Svenstrupgaard re-establishment centre, Jægergårdsgade drop-in centre, Reden (International, Copenhagen, Odense, Aarhus and Aalborg), Redernes Krisecenter and DanChurchSocial’s the Compass, Mariatjenesten, Hillerødgade, shelter Aalborg, shelter Aarhus, shelter Nørregade and shelter Østergade

---

**Supplementary table 1:** Baseline Characteristics of the cohort divided by seropositive and seronegative status.

|  | Seronegative | Seropositive | p |
| --- | --- | --- | --- |
| **n** | 764 | 55 |  |
| **PEH (%)** | 585 (76.6) | 43 (78.2) | 0.914 |
| **Age (median [IQR])** | 49.09 [38.89, 58.15] | 53.23 [45.90, 57.88] | 0.093 |
| **Gender (%)** |  |  | 0.650 |
| Female | 332 (43.5) | 27 (49.1) |  |
| Male | 416 (54.5) | 28 (50.9) |  |
| Other | 2 (0.3) | 0 (0.0) |  |
| NA | 14 (1.8) | 0 (0.0) |  |
| **Body mass index (mean (SD))** | 24.79 (4.94) | 27.66 (6.66) | <0.001 |
| **Smoker (%)** | 554 (72.5) | 33 (60.0) | 0.067 |
| **Alcohol use (%)** |  |  | 0.048 |
| Yes | 441 (57.7) | 32 (58.2) |  |
| NA | 31 (4.1) | 6 (10.9) |  |
| **Previously tested (%)** |  |  | 0.553 |
| Yes | 490 (64.1) | 35 (63.6) |  |
| NA | 22 (2.9) | 3 (5.5) |  |

PEH: people experiencing homelessness. Smoker: either previously or currently smoking. IV drugs: use of intravenous drugs. Smoked drugs: drugs that were reported smoked. Alcohol use: intake of alcohol within the past 12 months. Never COVID-19 Tested: never previously been tested for COVID-19.

**Supplementary table 2:** Symptoms stratified according to PEH and shelter workers.

|  | PEH | Shelter Worker | p |
| --- | --- | --- | --- |
| **n** | **628** | **191** |  |
| **Seropositive (%)** | 43 (6.8) | 12 (6.3) | 0.914 |
| **Any symptom (%)** | 303 (48.2) | 138 (72.3) | <0.001 |
| **Fever $\geq 38^{\circ}\text{C}$ (%)** | 116 (18.5) | 74 (38.7) | <0.001 |
| **Chills (%)** | 104 (16.6) | 48 (25.1) | 0.010 |
| **Loss of Smell (%)** | 50 (8.0) | 21 (11.0) | 0.247 |
| **Loss of Taste (%)** | 46 (7.3) | 15 (7.9) | 0.931 |
| **Sore Throat (%)** | 145 (23.1) | 105 (55.0) | <0.001 |
| **Cough (%)** | 229 (36.5) | 102 (53.4) | <0.001 |
| **Shortness of breath (%)** | 127 (20.2) | 35 (18.3) | 0.636 |
| **$\geq 3$ Symptoms (%)** | 142 (22.6) | 73 (38.2) | <0.001 |

Symptoms experienced since March 1<sup>st</sup> 2020. $\geq 3$ Symptoms; participants who registered three or more symptoms.

## References

1. World Health Organization. Coronavirus disease (COVID-19) pandemic [Internet]. [cited 2021 Mar 17]. Available from: https://www.who.int/emergencies/diseases/novel-coronavirus-2019

2. Danish Health Authority. COVID-19 Surveillance [Internet]. 2021 [cited 2021 Mar 17]. Available from: https://www.sst.dk/en/English/Corona-eng/Status-of-the-epidemic/COVID-19-updates-Statistics-and-charts

3. Platt L, Elmes J, Stevenson L, Holt V, Rolles S, Stuart R. Sex workers must not be forgotten in the COVID-19 response. The Lancet. 2020.

4. Danish Health Authority. General Guidance [Internet]. [cited 2021 Mar 17]. Available from: https://www.sst.dk/en/English/Corona-eng/Prevent-infection/General-guidance

5. Ralli M, Arcangeli A, Ercoli L. Homelessness and COVID-19: Leaving no one behind. Ann Glob Heal. 2021;87(1):1–3.

6. Benjaminsen L. Hjemløshed i Danmark 2019 National kortlaegning [Internet]. 2019 [cited 2021 Mar 17]. Available from: www.vive.dk

7. Folketal - Danmarks Statistik [Internet]. [cited 2021 Apr 26]. Available from: https://www.dst.dk/da/Statistik/emner/befolkning-og-valg/befolkning-og-befolkningsfremskrivning/folketal

8. Roederer T, Mollo B, Vincent C, Nikolay B, Llosa AE, Nesbitt R, et al. Seroprevalence and risk factors of exposure to COVID-19 in homeless people in Paris, France: a cross-sectional study. Lancet Public Heal. 2021;

9. Fazel S, Geddes JR, Kushel M. The health of homeless people in high-income countries: Descriptive epidemiology, health consequences, and clinical and policy recommendations. The Lancet. 2014.

10. Aldridge RW, Story A, Hwang SW, Nordentoft M, Luchenski SA, Hartwell G, et al. Morbidity and mortality in homeless individuals, prisoners, sex workers, and individuals with substance use disorders in high-income countries: a systematic review and meta-analysis. Lancet. 2018;

11. Christiansen, M L S., Moustgaard, H M., Lundgren, J., Rosholm J-U. Personer med øget risiko for et al vorligt sygdomsforløb med COVID-19. Ration Farmakoter [Internet]. 2020 [cited 2021 Mar 17];(7). Available from: https://www.sst.dk/da/Udgivelser/2020/Rationel-Farmakoterapi-7-2020/Personer-med-oeget-risiko-for-et-alvorligt-sygdomsforloeb-med-COVID-19

12. Danish Health Authority. Personer med øget risiko ved COVID-19. 2020.

13. Croxford S, Platt L, Hope VD, Cullen KJ, Parry J V., Ncube F. Sex work amongst people who inject drugs in England, Wales and Northern Ireland: Findings from a national survey of health harms and behaviours. Int J Drug Policy. 2015;

14. Shannon K, Crago AL, Baral SD, Bekker LG, Kerrigan D, Decker MR, et al. The global response and unmet actions for HIV and sex workers. The Lancet. 2018.

15. Grath-Lone LM, Marsh K, Hughes G, Ward H. The sexual health of female sex workers compared with other women in England: Analysis of cross-sectional data from genitourinary medicine clinics. Sex Transm Infect. 2014;

16. Ward H, Day S, Weber J. Risky business: Health and safety in the sex industry over a 9 year period. Sexually Transmitted Infections. 1999;

17. Statens Serum Institut. Vi Tester Danmark [Internet]. 2020 [cited 2021 Mar 17]. Available from: https://www.vitesterdanmark.dk/

18. CTK Biotech inc. OnSite COVID-19 IgG/IgM Rapid Test - (Serum / Plasma / Whole Blood). 2020.

19. Lassaunière R, Frische A, Harboe ZB, Nielsen ACY, Fomsgaard A, Krogfelt KA, et al. Evaluation of nine commercial SARS-CoV-2 immunoassays [Internet]. medRxiv. medRxiv; 2020 [cited 2021 Apr 26]. p. 2020.04.09.20056325. Available from: https://doi.org/10.1101/2020.04.09.20056325

20. Harris PA, Taylor R, Minor BL, Elliott V, Fernandez M, O’Neal L, et al. The REDCap consortium: Building an international community of software platform partners [Internet]. Vol. 95, Journal of Biomedical Informatics. Academic Press Inc.; 2019 [cited 2021 Apr 26]. Available from: https://doi.org/10.1016/j.jbi.2019.103208

21. Harris PA, Taylor R, Thielke R, Payne J, Gonzalez N, Conde JG. Research electronic data capture (REDCap)-A metadata-driven methodology and workflow process for providing translational research informatics support. J Biomed Inform [Internet]. 2009 Apr [cited 2021 Apr 26];42(2):377–81. Available from: /pmc/articles/PMC2700030/

22. Udmeldinger om alkohol - Sundhedsstyrelsen [Internet]. [cited 2021 Apr 26]. Available from: https://www.sst.dk/da/Viden/Alkohol/Alkoholforebyggelse/Sundhedsstyrelsens-dmeldinger-om-alkohol

23. ETHOS European Typology of Homelessness and Housing Exclusion. What is ETHOS [Internet]. [cited 2021 Mar 17]. Available from: https://www.feantsa.org/download/ethos2484215748748239888.pdf

24. FEANTSA. Working hard to end Homelessness in Europe [Internet]. [cited 2021 Mar 17]. Available from: https://www.feantsa.org/en

25. RStudio. Open source & professional software for data science teams - RStudio [Internet]. [cited 2021 Mar 17]. Available from: https://rstudio.com/

26. Baggett TP, Keyes H, Sporn N, Gaeta JM. Prevalence of SARS-CoV-2 Infection in Residents of a Large Homeless Shelter in Boston. JAMA - J Am Med Assoc. 2020;323(21):2191–2.

27. Mosites E, Parker EM, Clarke KEN, Gaeta JM, Baggett TP, Imbert E, et al. Assessment of SARS-CoV-2 Infection Prevalence in Homeless Shelters — Four U.S. Cities, March 27– April 15, 2020. MMWR Morb Mortal Wkly Rep [Internet]. 2020 May 1 [cited 2021 Apr 6];69(17):521–2. Available from: http://www.cdc.gov/mmwr/volumes/69/wr/mm6917e1.htm?s_cid=mm6917e1_w

28. Tobolowsky FA, Gonzales E, Self JL, Rao CY, Keating R, Marx GE, et al. COVID-19 Outbreak Among Three Affiliated Homeless Service Sites — King County, Washington, 2020. MMWR Morb Mortal Wkly Rep [Internet]. 2020 May 1 [cited 2021 Apr 6];69(17):523–6. Available from: /pmc/articles/PMC7206987/

29. UNAIDS. Sex workers must not be left behind in the response to COVID-19 [Internet]. 2020 [cited 2021 Mar 17]. Available from: https://www.unaids.org/en/resources/presscentre/pressreleaseandstatementarchive/2020/april/20200408_sex-workers-covid-19

30. Iversen K, Bundgaard H, Hasselbalch RB, Kristensen JH, Nielsen PB, Pries-Heje M, et al. Risk of COVID-19 in health-care workers in Denmark: an observational cohort study. Lancet Infect Dis [Internet]. 2020 Dec 1 [cited 2021 Mar 17];20(12):1401–8. Available from: www.thelancet.com/infection

31. Statens Serum Institut. Covid-19: Den Nationale Prævalensundersøgelse: Resultaterne fra 3. runde af antistofundersøgelse med 70.000 inviterede deltagere, uge 38-51, 2020 [Internet]. 2021 [cited 2021 Mar 17]. Available from: https://files.ssi.dk/praevalensundersoegelse_runde3

32. Laura Espenhain A, Tribler S, Svaerke Jørgensen C, Holm Hansen C, Wolff Sönksen U, Ethelberg S, et al. Title Prevalence of SARS-CoV-2 antibodies in Denmark 2020: results from nationwide, population-based sero-epidemiological surveys. medRxiv [Internet]. 2020 Apr 9 [cited 2021 Apr 26];2021.04.07.21254703. Available from: https://doi.org/10.1101/2021.04.07.21254703

33. Rogers JH, Link AC, McCulloch D, Brandstetter E, Newman KL, Jackson ML, et al. Characteristics of COVID-19 in Homeless Shelters : A Community-Based Surveillance Study. Ann Intern Med [Internet]. 2021 Jan 1 [cited 2021 Apr 6];174(1):42–9. Available from: /pmc/articles/PMC7517131/

34. Karb R, Samuels E, Vanjani R, Trimbur C, Napoli A. Homeless shelter characteristics and prevalence of SARS-CoV-2. West J Emerg Med [Internet]. 2020 [cited 2021 Apr 6];21(5):1048–53. Available from: /pmc/articles/PMC7514394/

35. Wang QQ, David •, Kaelber C, Xu • Rong, Volkow ND. COVID-19 risk and outcomes in patients with substance use disorders: analyses from electronic health records in the United States. Mol Psychiatry [Internet]. 2021 [cited 2021 Apr 6];26:30–9. Available from: https://doi.org/10.1038/s41380-020-00880-7

